# Neural networks for classification and image generation of aging in genetic syndromes

**DOI:** 10.1101/2021.12.09.21267472

**Authors:** Dat Duong, Ping Hu, Cedrik Tekendo-Ngongang, Suzanna Ledgister Hanchard, Simon Liu, Benjamin D. Solomon, Rebekah L. Waikel

**Affiliations:** Medical Genomics Unit, National Human Genome Research Institute, Bethesda, MD, United States of America

## Abstract

In medical genetics, one application of neural networks is the diagnosis of genetic diseases based on images of patient faces. While these applications have been validated in the literature with primarily pediatric subjects, it is not known whether these applications can accurately diagnose patients across a lifespan. We aimed to extend previous works to determine whether age plays a factor in facial diagnosis, as well as to explore other factors that may contribute to the overall diagnosis accuracy. To investigate this, we chose two relatively common conditions, Williams syndrome and 22q11.2 deletion syndrome. We built a neural network classifier trained on images of affected and unaffected individuals of different ages. Our classifier outperformed clinical geneticists at recognizing face images of these two conditions within each of the age groups (the performance varied between the age groups): (1) under 2 years old, (2) 2-9 years old, (3) 10-19 years old, (4) 20-34 years old, and (5) ≥35 years old. The overall accuracy improvement by our classifier over the clinical geneticists was 15.5% and 22.7% for Williams syndrome and 22q11.2 deletion syndrome, respectively. Additionally, comparison of saliency maps revealed that key facial features learned by the neural network differed slightly with respect to age. Finally, joint training real images with multiple different types of fake images created by a generative adversarial network showed up to 3.25% accuracy gain in classification accuracy.

## Introduction

Neural networks are emerging as powerful tools in many areas of biomedical research and are starting to impact clinical care. In the field of genomics, these methods are applied in multiple ways, including generating differential diagnoses for patients with possible genetic syndrome based on images,^1-4^ analysis of DNA sequencing data^5^ (including phenotype-based annotation^6^ and variant classification^7^), and prediction of protein structure.^8; 9^

In clinical genetics, the rarity, complexity, and number of different diseases,^10; 11^ coupled with a dearth of trained experts,^12; 13^ can lead to delayed diagnosis and suboptimal management.^14^ This can disproportionately impact older patients, as many clinical geneticists focus on pediatric diagnosis.^13^ Despite these challenges, previous large-scale applications of neural networks in clinical genetics studying individuals affected with many different genetic conditions have yielded impressive results.^1; 3^ In this paper, we endeavored to build upon these existing works by collating our own age annotated datasets to study the impact of age on facial diagnosis as well as to perform additional neural network analyses, which can also be extended to larger datasets or applied to different conditions.

We chose two distinct genetic conditions for further study: Williams syndrome (WS) (MIM 194050), which affects approximately 1 in 7,500 live births, and 22q11.2 deletion syndrome (22q), sometimes imperfectly referred to as “DiGeorge syndrome” (MIM 188400), which affects approximately 1 in 4,000-7,000 live births.^15-17^ We selected these conditions as they may be recognizable from facial features (in addition to other manifestations),^15; 16; 18; 19^ and based on relative data availability, which is still very limited compared to more common health conditions. Additionally, these two conditions represent varying ease of facial diagnosis: WS may have more consistently recognizable facial features, whereas 22q may have a more subtle facial presentation, which likely contributes to underdiagnosis for this as well as other conditions. To examine the influence of age on facial recognition, we evaluated how well clinical geneticists and our classifier recognize these conditions based on facial images of varying ages. We further explored additional neural networks applications to study both facial recognition as a whole and as a function of age. Our contributions can be summarized in the following four points.

**First**, we collected a dataset of publicly available WS and 22q images, which may be larger than others previous studied.^1; 3; 20^ Beyond the dataset, our approaches (and available code) may be used as subcomponents of other algorithms.^21^ We trained a neural network classifier on our dataset (N=1,894), which is still small compared to many deep learning datasets, thus pushing the capability of the neural network model. **Second**, we compared our classifier to clinical geneticists at recognizing these two syndromes for individuals in these five age brackets: infant; child; adolescent; young adult; older adult. **Third**, conditioned on WS or 22q, we calculated how much key facial features (analyzed via saliency maps) identified by the classifier differ with respect to age. This type of approach is important for DL in biomedical contexts,^22^ including related to disease progression and other temporal factors. **Fourth**, we estimated the effect of joint training real images with fake images created by a generative adversarial network (GAN). We evaluated four different types of GAN-based images: (1) unique faces for a specific disease and age group; (2) similar faces (e.g., related to skin tone or hair color) at different ages for a specific disease; (3) the same face at different ages for a specific disease; (4) faces containing characteristics of two different diseases (e.g. a hypothetical person having WS and 22q), used to determine if this could improve the classifier’s accuracy in differentiating images.

## Materials and Methods

### Ethics review

The study was reviewed by National Human Genome Research Institute (NHGRI) bioethicists and the National Institutes of Health (NIH) Institutional Review Board (IRB). The main analyses were considered not human subjects research; a waiver of consent was granted by the NIH IRB (NIH protocol: 000537) for the work involving the surveys of medical professionals, as described below.

### Data collection

We searched Google and PubMed using the disease names of interest to select publicly available images depicting individuals with WS, 22q, or other genetic conditions that may resemble WS or 22q (see Table S1 for more details about these conditions). Below, when the context is clear, we will refer to these “other genetic conditions” as the control group. From the available source information for each image, we categorized the images into five age brackets: (1) infant (under 2 years old), (2) child (2-9 years old), (3) adolescent (10-19 years old), (4) young adult (20-34 years old), and (5) older adult (≥35 years old). We attempted to collect images of individuals from diverse ancestral backgrounds, though standardized and complete information regarding race and ethnicity was often unavailable (Table S2). In total, we collected 1,894 images, and partitioned them into 1,713 and 181 train and test images, respectively (Table S3). Image sets included both color and black and white images, with varying image resolution. Test images were selected from color images subjectively judged to have adequate resolution for human viewing, and which included representations of both sexes and of apparently ancestrally diverse individuals, though we recognize the many challenges in these and related areas.^23^ The control group test images included individuals with other genetic and congenital conditions, including those with overlapping facial features with WS or 22q. We applied StyleGAN face detector and image preprocessing to rotate and center our images, and manually aligned images that failed this preprocessing step.

### Classifier

We selected the EfficientNet-B4 classifier, which obtained high performance on the ImageNet data with a relatively low number of parameters.^24^ We loaded the weights pretrained on ImageNet and continued training EfficientNet-B4 end-to-end. Combining and then jointly training a small dataset of interest with a larger auxiliary dataset often increases the prediction accuracy.^25; 26^ Our auxiliary dataset is the FairFace dataset, which contains 108,000 “in-the-wild” faces (i.e., faces oriented in various angles and/or partially covered with hands, hats or sunglasses) of equal ratio from (using definitions in FairFace) white, black, Latino, East/Southeast Asian, Indian, and Middle Eastern populations.^27^

StyleGAN face detector and image preprocessing, such as rotating and centering faces, were applied to the in-the-wild FairFace faces, resulting in 62,088 usable images. We partitioned these 62,088 images into the age groups described above via FairFace age classifier. One sixth of the images (N=10,348) in each age category was randomly chosen as test images, which we evaluated with our own test images. The remaining 51,740 images were used with our images to train EfficientNet-B4.

Because of our small data set and the assumption that at least some features persisted across age groups, we trained EfficientNet-B4 on our images (regardless of age), and FairFace images, to recognize the 4 labels: WS, 22q, other genetic conditions (control), and unaffected. We included unaffected individuals, as an important consideration in clinical practice is the ability to differentiate a potentially affected from an unaffected person, especially as some genetic conditions can have subtle findings often missed by general clinicians as well as subspecialists. The classifier was trained with cross entropy loss function where 1-hot encodings represent true image labels. We rescaled all the images into resolution 448 × 448 pixels when training EfficientNet-B4. Image resolution was chosen to maximize GPU usage (two Nvidia P100, training batch size 64).

We trained 5 classifiers via 5-fold cross-validation (one for each fold), and then created an ensemble predictor by averaging the predicted label probabilities of an image from these 5 classifiers. When averaging, we considered only the classifiers that produced a maximum predicted probability (over all the labels) of at least 0.5. Our code is available at https://github.com/datduong/Classify-WS-22q-Img.

### Comparison to clinicians

We compared our classifier to board-certified or board-eligible clinical geneticist physicians via surveys sent by Qualtrics (Provo, Utah, United States). As WS and 22q syndromes are relatively distinct, we felt that it was more meaningful to evaluate WS test images against their own controls, and likewise for 22q test images. We emphasize that, for nontrivial comparisons, the control test images were of conditions resembling WS and 22q. For WS surveys, there were 50 WS (10 images per age group) and 50 corresponding control test images. To keep survey length reasonable, each participant went through a random subset of 25 WS (5 images per age group) and 25 control images. The ordering of the selected images in a survey was randomized, and the answer choice for a question was either “Williams Syndrome” or “Other Condition”. The same setup was also employed for 22q surveys. In addition to asking clinical geneticists to classify images, we also asked questions about the impact of patients’ age on diagnosis to determine attitudes and opinions on the age in the diagnosis process. See Supplemental information or https://github.com/datduong/Classify-WS-22q-Img for example surveys.

Following previous methods,^21; 28; 29^ we estimated that 30 participants would provide a statistical power of 95% to detect a 10% difference. Participants were recruited via email. To identify survey respondents, we obtained email addresses through professional networks, departmental websites, journal publications, and other web-available lists. A total of 225 clinical geneticists were contacted, 36 completed the 22q survey and 34 completed the WS survey. If multiple respondents completed the same survey, only the first survey was used for analysis. See Table S4 for a description of survey respondents.

### Generative adversarial network (GAN)

We trained a GAN for each data partition from the 5-fold cross-validation in section “*Classifier”*. We describe the GAN training and image generation for a data partition *p*, which also will apply to the other partitions. Partition *p* contains our images of affected individuals and FairFace unaffected individuals. Ideally, we would train GAN on all these individuals. This may blend the features of different racial/ethnic groups in FairFace with our dataset, and the GAN would generate diverse images of affected individuals. However, a partition *p* approximately has 41,392 images of unaffected FairFace individuals, and our preliminary GAN experiments required a large amount of computational power. The larger FairFace dataset also often skewed GAN output where the generated images of affected individuals looked more like the unaffected subset. Therefore, in a partition *p*, we trained GAN on our images and a fixed subset of FairFace. This subset was randomly chosen with 500 individuals in each age bracket. In each training batch, we selected an equal number of affected and unaffected individuals.

Our GAN is based on the conditional StyleGAN2-Ada and generates images using both disease statuses and age categories.^30^ We made the following key modification to StyleGAN2-Ada. The default label embedding is L × 512, where L is the number of labels, and produces a vector of length 512 for each label. Training this embedding requires many people with a specific disease in a certain age category. However, some disease and age label combinations have small sample sizes; for example, our dataset has 39 WS and 35 22q individuals older than 35 years of age. We replaced the default label embedding with two smaller 4 × 256 and 5 × 256 matrices to represent the 4 diseases (WS, 22q, other conditions, and unaffected) and 5 age categories. Training the 4 × 256 disease embedding then uses all affected individuals in every age category. Likewise, training the 5 × 256 age embedding uses all the images in our dataset and in FairFace. The outputs of these two components are concatenated to a vector of size 512 to match the rest of the StyleGAN2-Ada architecture. Hence, except for the label embeddings, we initialized all the StyleGAN2-Ada weights with the pretrained values on FFHQ dataset at resolution 256 × 256 pixels;^30^ all fake images were also generated in the resolution 256 × 256 pixels. Image resolution was chosen to maximize GPU usage (two Nvidia P100).

After training GAN on a data partition *p*, we generated four types of fake images: (1) unrelated faces for a specific disease and age group; (2) similar faces at different ages for a specific disease; (3) the same face at different ages for a specific disease; (4) faces containing characteristics of two different conditions, which we hypothesized could aid classifier accuracy.

Because there are already many unaffected people in FairFace, we generated just images of affected individuals from the disease label *d* ∈{WS, 22q, other condition} and age bracket *a* ∈{infant, child, adolescent, young adult, older adult}. The number of generated images for each pair (*d,a*) is equal to the average count of all age groups with a specific disease *d* in the data partition. Thus, in a specific disease, respectively we made more and fewer images for the uncommon and common age groups. In total, each image type has the same count as the size of the affected individuals in the data partition in which the GAN was trained.

For type 1, we generated a fake image *i* by concatenating the random vector *r*_*iad*_ with the label embedding *e*_*d*_ and *e*_*a*_, denoted as [*r*_*iad*,_ *e*_*a*,_ *e*_*d*_], and then passing this new vector to our GAN image generator. Each disease *d* and age group *a* combination has images generated from their own unique random vector *r*_*iad*_ so that all the fake images are theoretically unique (Figure 1a).

**Figure 1.**
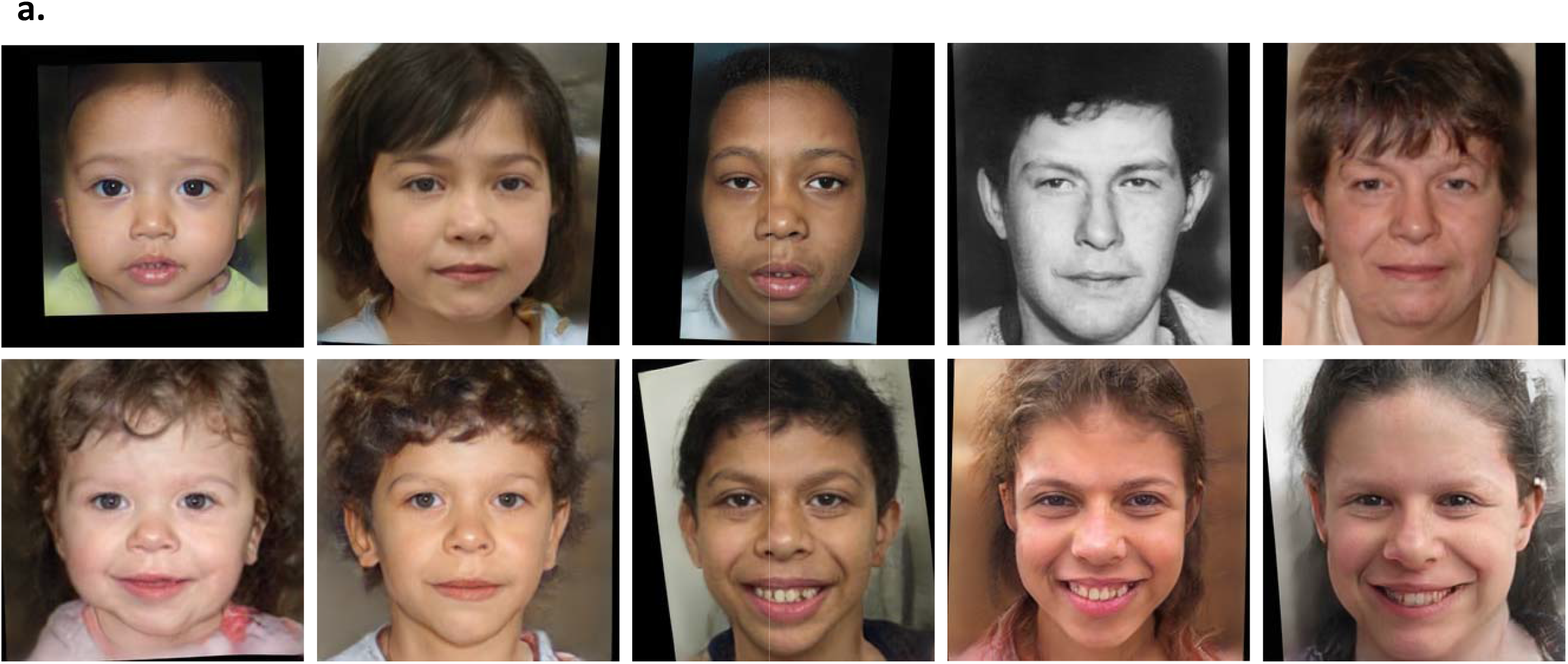
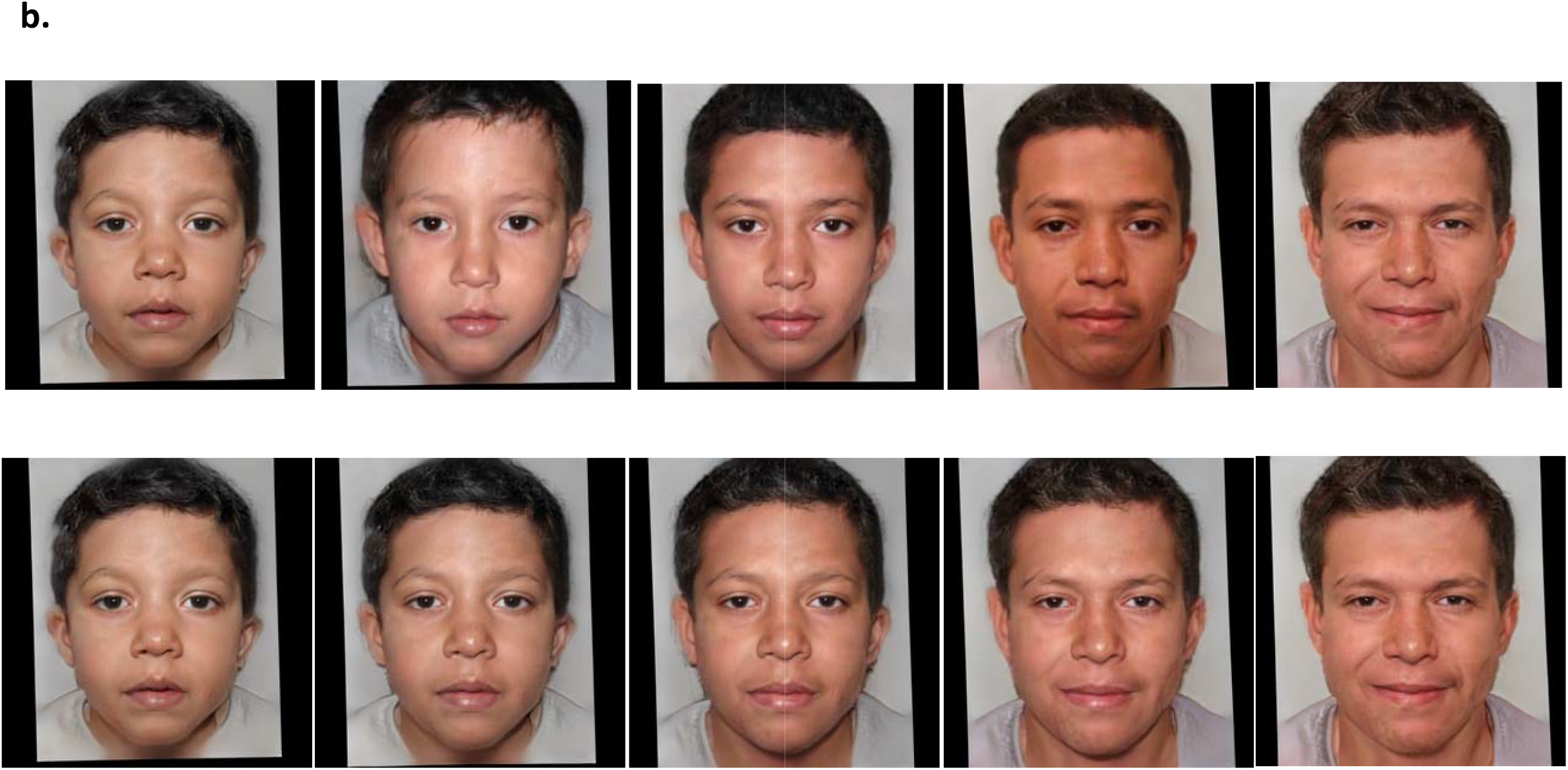
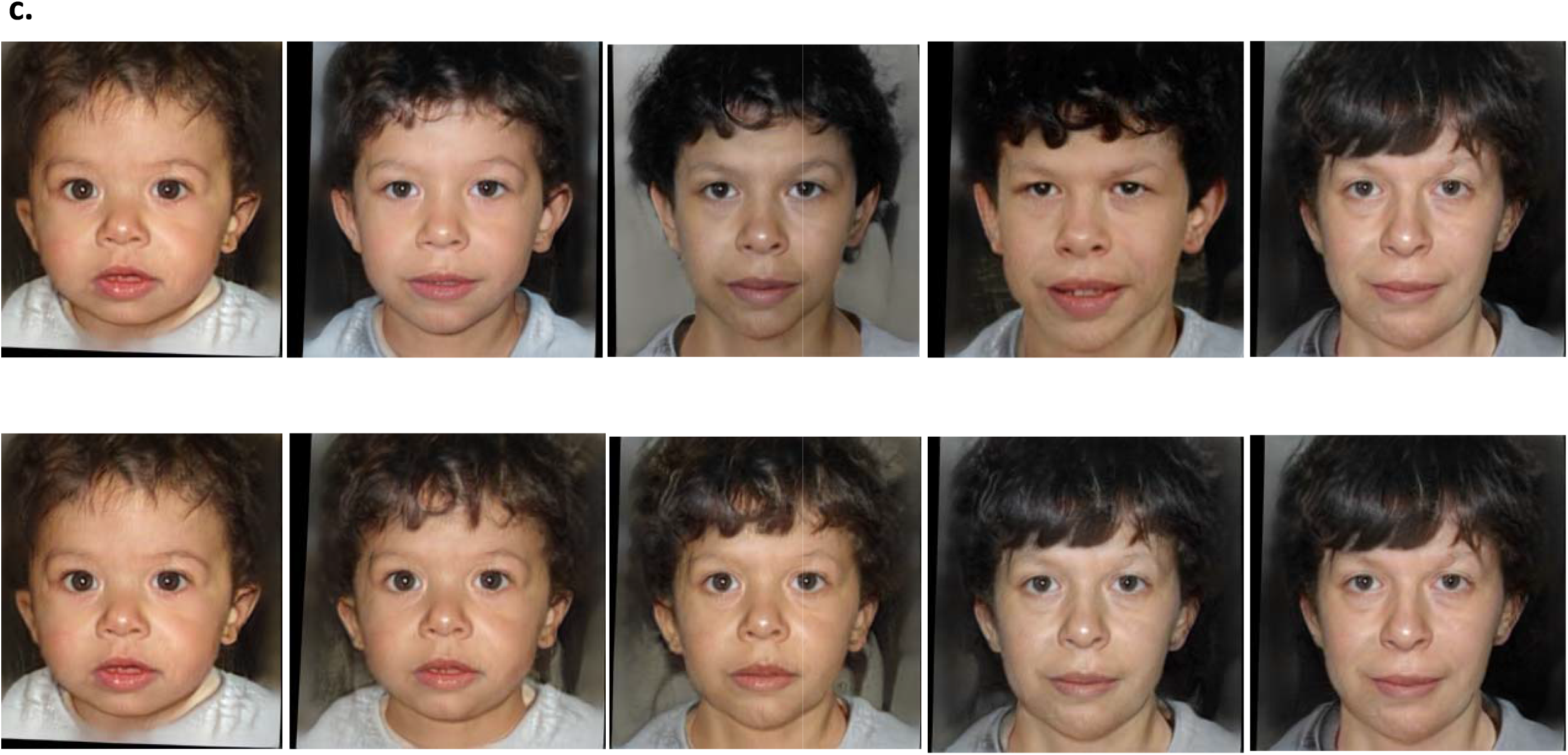
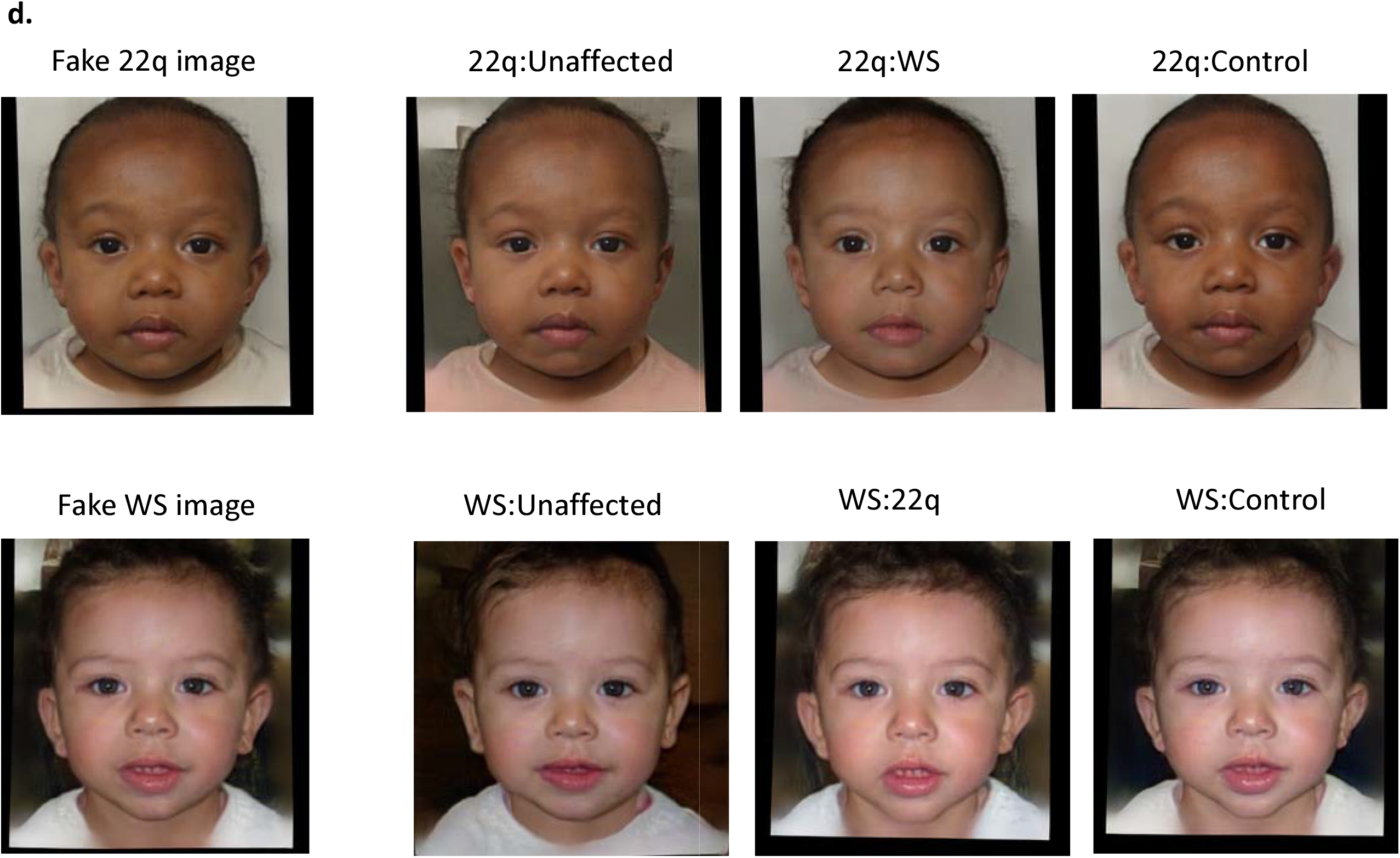
Examples of GAN fake images for 22q and WS. Type 1 fake images **(a)** 22q (top row) and WS (bottom row) were generated with GAN and are all theoretically unique. Type 2 (top row) and type 3 (bottom row) fake images 22q **(b)** and WS **(c)** were generated using the same random vector in each age group. For type 2, general features, such as skin tone and hair color, are roughly preserved. For type 3, the images look consistent at depicting the same “person” progresses through different age groups. Type 4 fake images **(d)** were created images with blended facial characteristics of two disease labels. The main disease condition (22q or WS) represents 55% of the facial phenotype and the added disease condition represents 45% of the facial phenotype. For example, WS:Unaffected is a blend of 55% WS facial features and 45% unaffected facial features. Only the blended images were used for training, the left most images are shown here as references.

Type 2 images are generated by varying the age embedding *e*_*a*_, where *a* ∈{infant, child, adolescent, young adult, older adult}, while fixing the random vector *r*_*id*_ and the disease embedding *e*_*d*_. The vector *r*_*id*_ is unique to the *i*^th^ image having disease *d*. Following from this, every disease has own unique images, but within the same disease the images at each age category have similar facial features such as skin tone and hair color (Figure 1b and 1c).

For type 3 images, we interpolated 3 equally spaced vectors between the age embedding e_*infant*_ and e_*older adult*_. For a disease *d*, we generated five fake images from a random vector *r*_*id*_ by passing into GAN generator these five inputs [*r*_*id*_, *e*_*infant*_, *e*_*d*_], [*r*_*id*_, *0*.*75e*_*infant*_+*0*.*25e*_*older adult*_, *e*_*d*_], [*r*_*id*_, *0*.*5e*_*infant*_*+0*.*5e*_*older adult*_, *e*_*d*_], [*r*_*id*_, *0*.*25e*_*infant*_ *+0*.*75e*_*older adult*_, *e*_*d*_], and [*r*_*id*_, *e*_*older adult*_, *e*_*d*_]. These images closely represent the same person affected with disease *d* at five different age groups (Figure 1b and 1c). There are additional potential approaches for depicting age progression, which we may explore in future studies.^31^ Of note, previous work^1; 3^ used different and/or additional age brackets, some of which may not involve sufficient numbers of images for robust analyses, at least in our datasets.

Type 4 images were generated like type 3; however, we reversed the roles of disease *d* and age label *a*. With a random vector *r*_*ia*_, we generated three fake images from the inputs [*r*_*ia*_, *e*_*a*_, *ce*_*WS*_ *+ (1-c)e*_*22q*_],[*r*_*ia*_, *e*_*a*_, *ce*_*WS*_ *+ (1-c)e*_*control*_], [*r*_*ia*_, *e*_*a*_, *ce*_*WS*_ *+ (1-c)e*_*unaffected*_] where *c* is a pre-defined fraction between 0 and 1. These images represent a person at age *a* having facial characteristics of two different diseases (Figure 1d). The true labels for type 4 images are soft labels; for example, the image created from the vector [*r*_*ia*_, *e*_*a*_, *ce*_*WS*_ *+ (1-c)e*_*22q*_] would have the label encoding [*c, 1-c, 0, 0*] instead of the traditional 1-hot encoding. Here, the training loss function is still cross-entry but for soft label.

Next, we created 4 new larger datasets by combining partition *p* with each of the 4 fake image types. We then trained EfficientNet-B4 on each of these new larger datasets. For each type of new dataset, we created the ensemble predictor over all the data partitions following the approach mentioned in section *“Classifier”*. Our code is available at https://github.com/datduong/stylegan2-ada-Ws-22q.

### Attribution analysis for features in different age groups

To visualize which facial features of an image the classifier considered to be important, we produced saliency maps using window size 20 × 20 pixels and stride 10 x10 pixels using the occlusion attribution method.^32^ For a test image, we averaged the saliency maps of the classifiers in the ensemble predictor. We used the permutation test to measure how much these facial features identified by the classifiers differ with respect to age. Our Qualtrics surveys had 10 test images in each disease and age label combination. Conditioned on a disease and two age groups, we permuted the 20 images into two sets and repeated this permutation 100 times. Each time, we averaged the saliency map over the 10 images in each set, and then retrieved the embedding of this average attribution via the EfficientNet-B4 trained on ImageNet. In each permutation, we computed the Euclidean distance between the embeddings of these 2 sets. If the observed Euclidean distance is smaller than 5% (or some other threshold) the permutation values, then the two age groups of a specific disease were defined as not statistically different. That is, the key facial features identified by the classifier do not differ with respect to age.

## Results

### Classifier accuracy

Our classifier, which was trained on images of individuals with WS, 22q, other genetic conditions, and those who are presumably unaffected, could correctly classify unique test images 68% to 100% of the time, with the lowest accuracy for 22q and the highest accuracy for unaffected individuals (Figure 2). While unaffected individuals are not misclassified as affected, the opposite is not typically true, presumably as some affected individuals may show only subtle features of the condition. Classification accuracy of WS (86%) was the highest among the affected individuals we examined; our results suggest that these individuals often clearly display key findings (e.g., the morphology of the eyes and mouth) compared to individuals with 22q, who are frequently misclassified (30%) as the control group (conditions other than WS or 22q).

**Figure 2.**
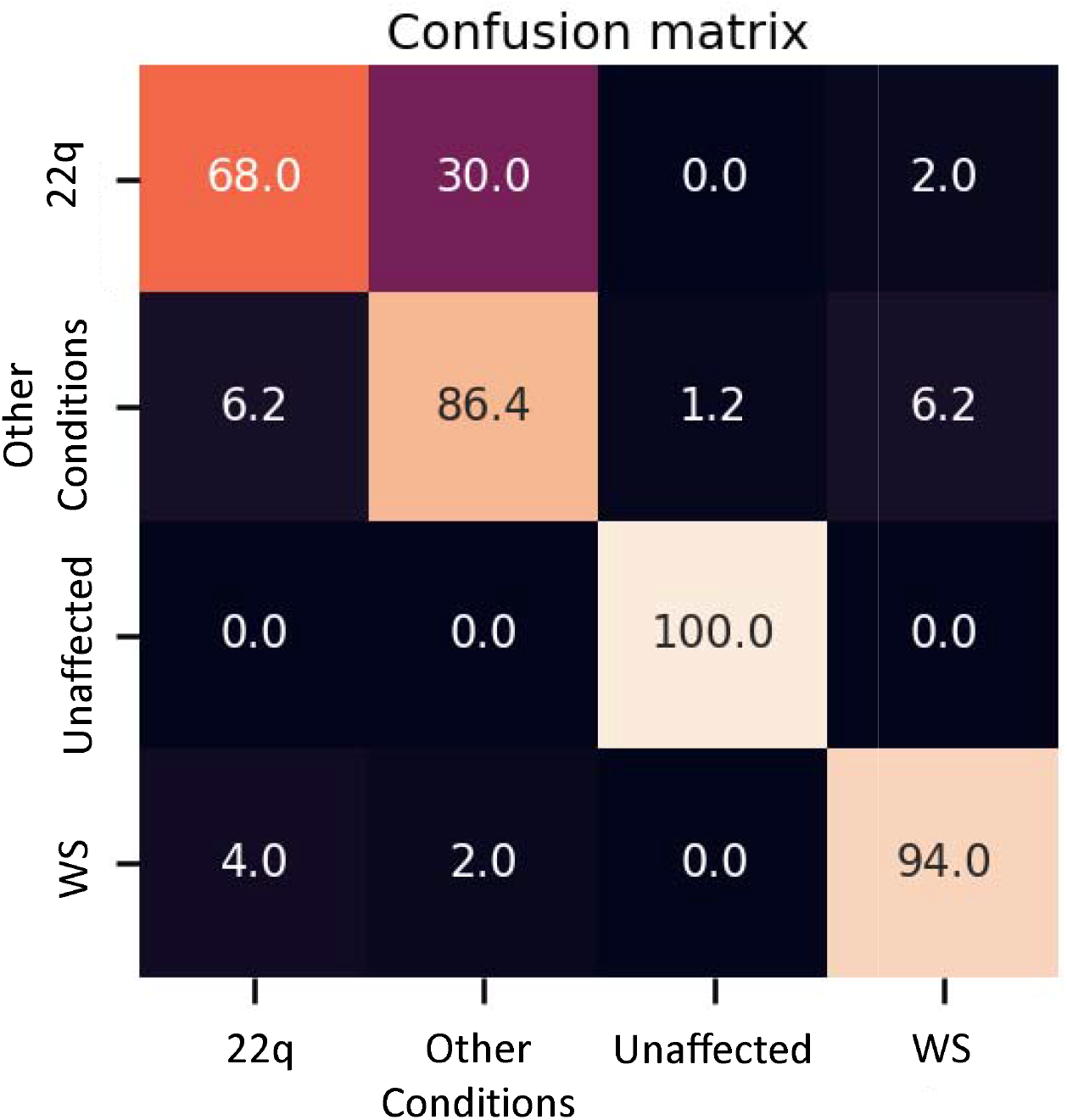
Confusion matrix of accuracy of classifier trained on real images. Rows represent the correct label, while columns represent the label chosen by the classifier. The diagonal numbers represent the percent accuracy for each category (the percentage of images when the correct label was identified), while the off-diagonal represent misclassification percentage ascribed to an incorrect category. Accuracy is based on 50 test images of WS, 50 of 22q and 81 of other conditions.

### Comparison to clinical geneticists

Clinical geneticists completed survey(s) classifying images of individuals with WS versus other conditions and 22q versus other conditions. These were two separate surveys as, based on preliminary testing, we felt that WS and 22q would typically be considered distinct by clinical geneticists, and that it would be more meaningful to evaluate these conditions separately. The statistical differences between clinical geneticists and our model were measured via paired t-test. As our model was trained with 4 labels, we took the highest prediction probability between WS and control for a test image in surveys containing WS and control images. The same idea applied to surveys containing 22q and their corresponding controls. Our model outperformed clinical geneticists by 15.5% (77.5% vs 93%, p = 6.828e^-11^) for WS and 22.7% (59.3% vs 82%, p = 3.203e^-13^) for 22q.

To determine whether age of patient affects accuracy, we determined the average accuracy for each age group (Table 1). On average, clinical geneticists had the most difficulty identifying infants affected with WS (67.3%) and the greatest accuracy with adolescents (80.7%). Clinical geneticists had the most difficulty classifying older adults with 22q (50.7%) and the greatest accuracy classifying adolescents (67.3%). Our classifier outperformed clinical geneticists in all age groups (Table S5). However, we emphasize that because each age group has 10 images, performance differences may represent only a few images. Despite the small test size per condition and age bracket, our results suggest clinical diagnosis may be more difficult in some age groups.

**Table 1.** Average accuracy over all 10 test images in each age group. Each test image was viewed and classified by 15 clinical geneticists. Our classifier, either trained on real images alone or on both real and GAN age progression (age prog) images, obtains higher accuracy for each age group, except for the oldest 22q cohort. P-values in parentheses comparing human against model were computed via permutation test.

|  | Age | 22q |  |  | WS |  |  |
| --- | --- | --- | --- | --- | --- | --- | --- |
|  |  | Human | Model |  | Human | Model |  |
|  |  |  | Real image | + Age prog |  | Real image | + Age prog |
| Disease | Infant | 0.54 | 0.7 (0.18) | 0.8 (0.05) | 0.673 | 1 (0) | 1 (0) |
|  | Child | 0.527 | 0.8 (0.04) | 0.9 (0) | 0.707 | 1 (0) | 1 (0) |
|  | Adolescent | 0.673 | 0.7 (0.44) | 0.8 (0.21) | 0.807 | 0.9 (0.24) | 1 (0) |
|  | Young Adult | 0.613 | 0.8 (0.10) | 0.9 (0.01) | 0.74 | 1 (0) | 1 (0) |
|  | Older Adult | 0.507 | 0.5 (0.50) | 0.5 (0.50) | 0.713 | 0.9 (0.09) | 1 (0) |
| Other Conditions | Infant | 0.753 | 1 (0) | 1 (0) | 0.84 | 0.9 (0.35) | 0.9 (0.35) |
|  | Child | 0.6 | 0.9 (0) | 0.9 (0) | 0.767 | 0.8 (0.40) | 0.8 (0.40) |
|  | Adolescent | 0.52 | 1 (0) | 1 (0) | 0.86 | 0.9 (0.39) | 0.9 (0.39) |
|  | Young Adult | 0.6 | 0.9 (0.01) | 1 (0) | 0.787 | 0.9 (0.19) | 0.9 (0.19) |
|  | Older Adult | 0.593 | 0.9 (0.01) | 0.9 (0.01) | 0.86 | 1 (0) | 1 (0) |

### Estimating facial differences across age groups

Conditioned on a disease, we compared the saliency maps of the test images in different age groups based on method in “*Attribution analysis for features in different age groups”*. Composite images of saliency maps averaged over all test images (occlusion analysis) were generated for each age group (Figure 3). Key facial features were identified by the classifier and are statistically different with respect to age assuming the standard 5% statistical significant threshold (Figure 4). For example, the observed Euclidean distance between the embeddings of the infant and child composite saliency maps (Figure 4) ranks higher than 43 of the 100 permutation values. The greatest differences are seen for the infant and older adults in both WS and 22q. Compared to 22q, WS facial features identified by our model differ more with respect to age, which may show how facial features of a syndrome can be age-specific. Again, we emphasize that there can be confounding factors. For example, a person’s facial expression (e.g., whether a person is smiling) may explain why features of adolescent WS test images are different than the other age groups (Figure 4). Clinical geneticists may also rely on non-specific facial clues to classify syndromes and conditions. For example, high sociability and friendliness are common features of WS.^18; 33^ While we did not intentionally select images based on facial expression (smiling or not smiling), we found that more WS test images (60%) had a partial or full smile compared to other conditions (44%). Clinical geneticists were more likely to misclassify a Williams syndrome image if not smiling (58.3%) vs. smiling (82.4%) (Table S6). The presence or absence of a smile did not impact classification of other conditions.

**Figure 3.**
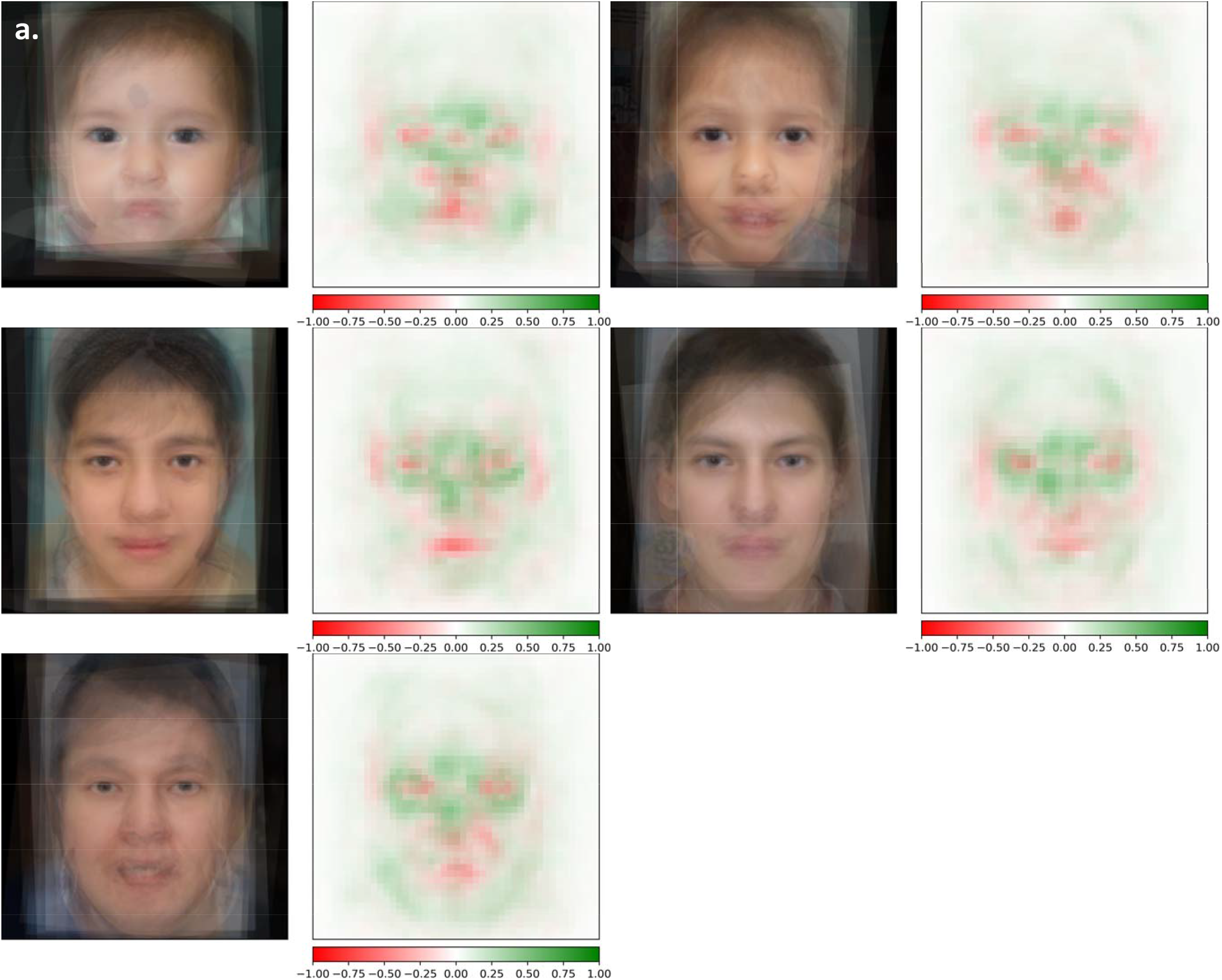
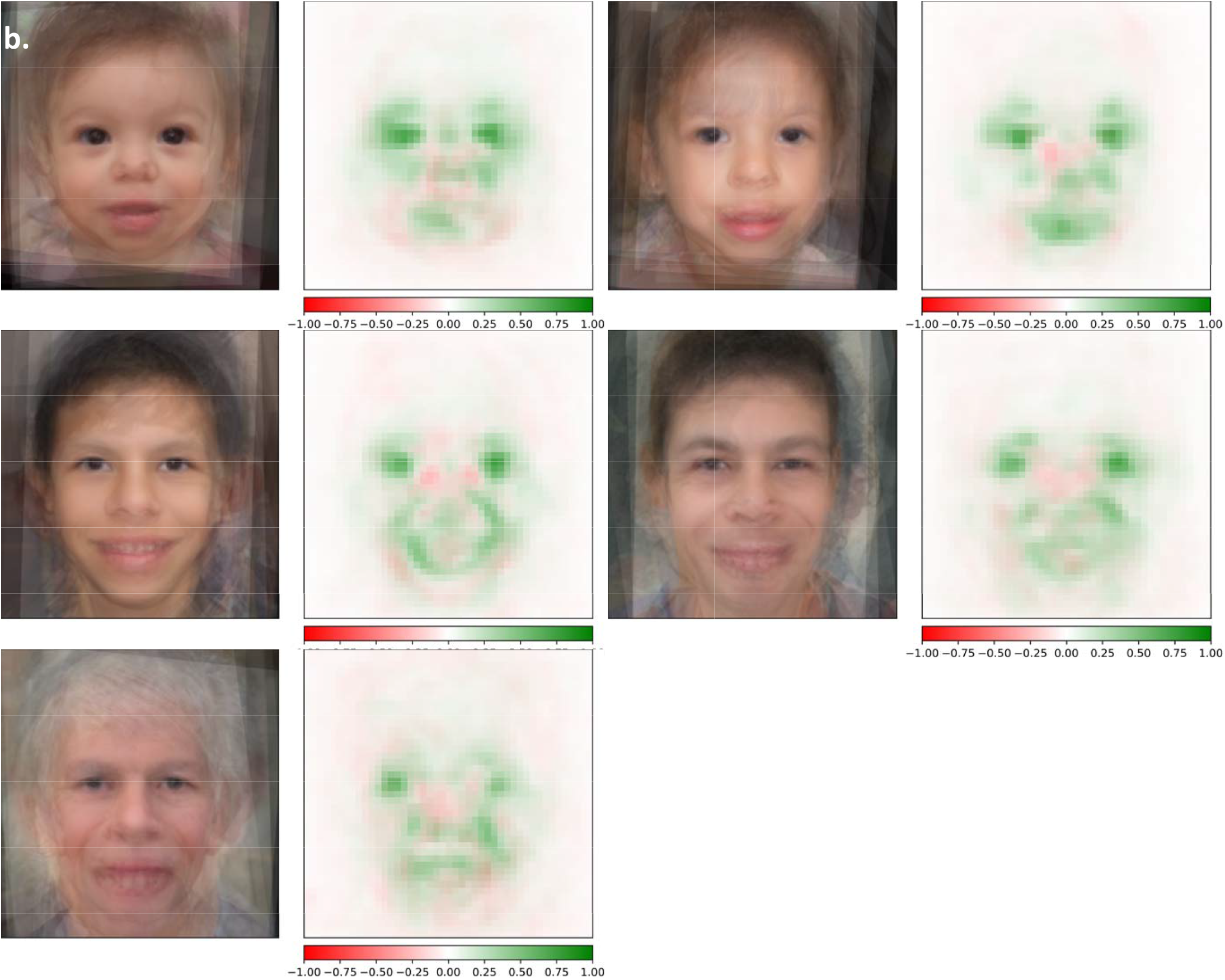
Visual analysis of 22q and WS facial images across the lifespan. Composite saliency maps (by occlusion method) were generated by averaging all test images in each age groups: infant, child, adolescent, young adult, and older adult (reading left to right) for both 22q **(a)** and WS **(b)**. Green and red indicate positive and negative contribution to the correct labels, respectively.

**Figure 4.**
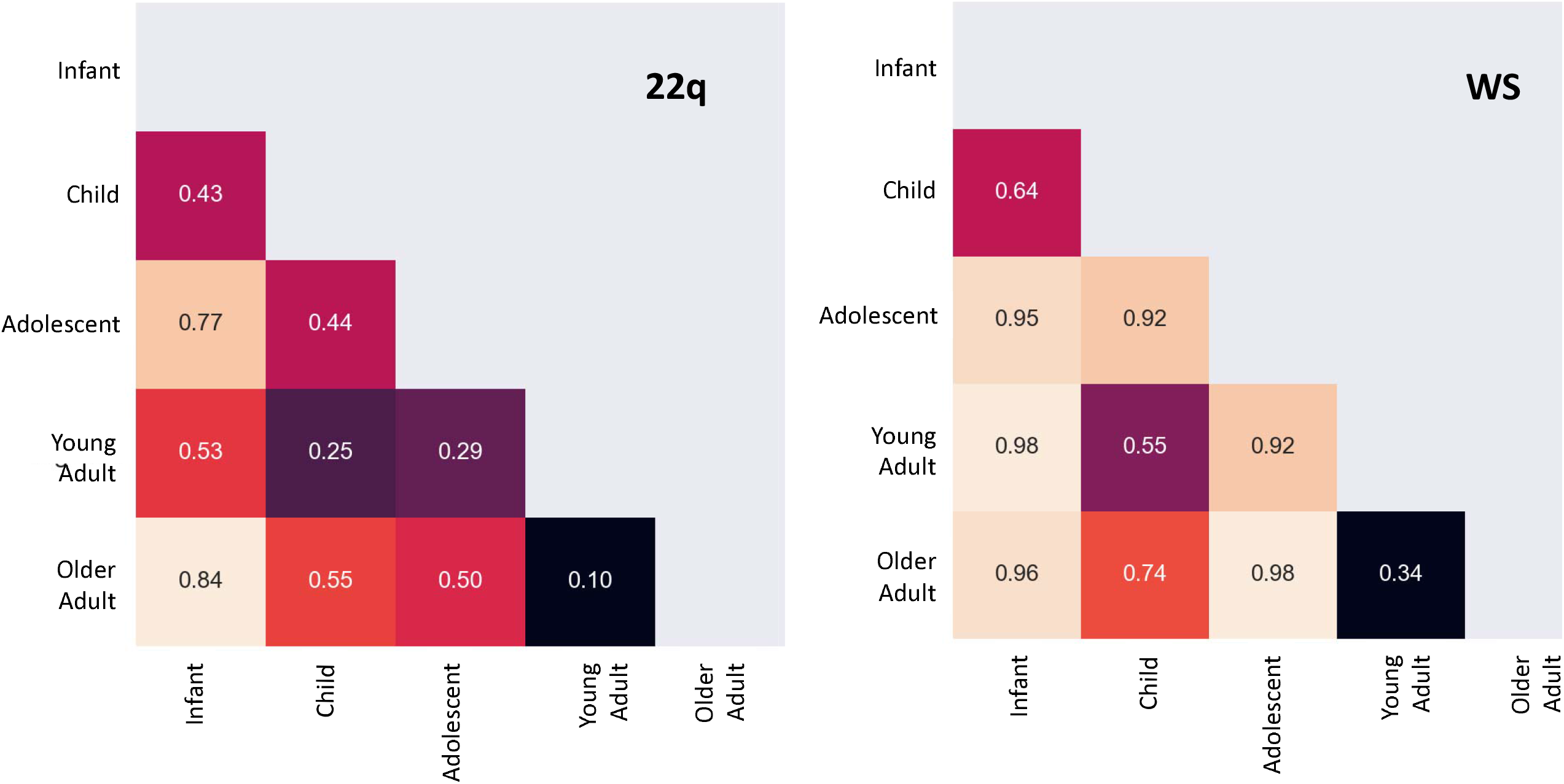
Rank of Euclidean distance matrix of key facial features during aging. Rank (in fraction) of observed Euclidean distance (out of 100 permutations) between embeddings of the averages of saliency maps for two age groups. A small number indicates key features identified by the neural network for two age groups are more statistically similar, whereas a larger number indicates key features are more statistically different. For example, there is more statistical difference between key facial features of an infant and adolescent (0.95) versus an infant and child with WS (0.64).

### Classifiers trained on real and GAN images

Table 2 shows the accuracies of the classifiers trained on real images and different types of GAN images (see Supplementary materials for the corresponding confusion matrices). Although the improvements are minimal, all four types of GAN images obtain slightly higher average accuracy compared to the base classifier, with type 3 GAN images (showing age progression for the same person) performing the best.

**Table 2.** Accuracy of classifier trained on real images compared to classifiers trained on both real images and each of the four types of fake images. Column names unrelated, age related, age progression (age prog), and blended correspond to fake GAN images of type 1, 2, 3, and 4, respectively. The greatest improvement in accuracy was observed with the addition of age progression fake images.

|  | Real images | + unrelated | + age related | + age prog | + blended 55-45% | + blended 75-25% |
| --- | --- | --- | --- | --- | --- | --- |
| **22q** | 68 | 76 | 74 | **76** | 68 | 76 |
| **Controls** | **86.4** | 81.5 | 82.7 | 85.2 | 85.2 | 85.2 |
| **Unaffected** | 100 | 100 | 100 | **100** | 100 | 100 |
| **WS** | 94 | 96 | 96 | **100** | 96 | 96 |
| **Average** | 87.1 | 88.375 | 88.175 | **90.35** | 87.3 | 89.3 |

We also compared the classifier trained with type 3 GAN images against the clinical geneticists in each age bracket (Table 1). The cumulative improvements over humans are 95% vs. 77.5% (p = 1.846e^-11^) for WS and 87% vs. 59.3% (p = 1.703e^-15^) for 22q.

We suspect that type 3 GAN images improved the base classifier more because key facial features varied with respect to age (Figures 3 and 4). By conditioning on the same person, we may capture finer details of how a disease progresses with time.

## Discussion

The practice of medical genetics has shifted considerably in the last several decades. One major reason is the growing availability of high-throughput genetic/genomic testing. This allows more precise diagnosis, and has changed the approach to phenotyping.^34^ However, access to these testing technologies is uneven, and it remains important to be able to quickly recognize patients who may be affected by certain conditions, especially those with near-term management implications.^11; 35^ For example, people with WS are prone to infantile electrolyte abnormalities and immunologic dysfunction, and people with 22q may be affected by endocrine, immunologic, cardiovascular, and other sequelae that require immediate attention.^18; 19^

To provide examples of ways to bolster the standard diagnostic process as well as to build on the impressive findings of previous, related studies,^1; 3; 4^ we analyzed and provide a larger dataset of WS and 22q individuals (although these other studies contained a much larger total number of individuals having multiple other diseases). We also compared results for different ages of individuals. Our classifier outperformed clinical geneticists at identifying WS and 22q individuals by large margins (15.5% and 22.7%) respectively. This was consistently true for each age group.

We hypothesized that because geneticists overall often have more clinical experience with children, and as the medical literature tends to focus more on pediatric presentations of congenital disorders, respondents would feel most confident about diagnosis in younger age groups and would also perform best with images of younger patients. However, for WS, our results show that respondents’ accuracy did not correlate with their confidence level in diagnosing the conditions at various ages. For example, 46.7% (14/30) and 50% (15/30) of clinical geneticists surveyed reported that infants and older adults with WS are difficult to classify based on facial features, respectively, but were able to classify these patients with similar accuracy to those of other ages (Table S7). This may imply other explanations. For example, clinicians may feel the most confident considering patients at ages they most often encounter, but this may not also be reflected in performance. Features of WS may also be more pronounced with age such that clinicians can more readily recognize the condition even with less experience. On the other hand, 60% (18/30) and 40% (12/30) of clinical geneticists, respectively, reported that infants and older adults affected with 22q are difficult to classify based on facial features, which aligns better with their performance. There are again multiple explanations, but one possibility is that 22q may simply be a more subtle condition, or that features do not become more obvious with age. To explore this and other questions further, we plan to extend our analyses in the future to additional images and conditions, including by determining which particular features are objectively assessed by humans. This may help reveal underlying reasons for diagnostic patterns.

Intuitively, due to sample size difference, a classifier trained on fake and real images should outperform the one trained on just real images. Interestingly, this approach does not always improve the prediction outcome in previous works from other disciplines.^36; 37^ Our result also showed that there was a small improvement (up to 3.25% accuracy gain). In the future, we plan to evaluate whether GAN images may be useful in other applications. For example, generated images could help as educational tools, or to generate realistic images to obviate data sharing and privacy concerns. Along these lines, our results also suggest areas of weakness that could be targeted for the generation of GANs, such as images of infants and older individuals or medical training purposes. It would also be informative to conduct a meta-analysis on existing literature across different disciplines to estimate the improvement of training GAN images and real images.

Our study has limitations. First, our dataset is small compared with other datasets for image recognition and may involve biases. Since collecting publicly free images of confirmed cases is challenging, we did not have balanced numbers of images for each condition and age bracket combination (Table S3), and the types of images may have differed in certain categories. For example, we included some gray-scale images; having different numbers of these in some subsets could affect the color consistency for GAN-based transformation (Figure S1). However, the average age for each age grouping was consistent (Table S8). For example, the average age for the child age grouping was 5.54, 5.01, and 5.33 years old for 22q, WS, and controls, respectively. Second, but also related to our sample size, we may have suboptimal grouping. For example, grouping all individuals older than a certain age into the oldest age group may have obscured differences within that group.

Despite the rarity (and therefore lack of data availability) of many genetic conditions, neural networks have high potential in the field, both due to the ability to accurately categorize patients based on underlying molecular causes, as well the lack of trained experts throughout the world such that these tools could be highly valuable.^38^ This area provides a ripe opportunity for patients, clinicians, researchers, and others to collaborate for the good of the impacted community. Privacy and data handling issues must be taken seriously; we hope that obstacles around data and code sharing can be addressed so as not to impose undue barriers to helping affected individuals and families.

## Data Availability

Data are available upon reasonable request, and will be publicly release upon final publication. Code to classify and generate patient images are currently available at https://github.com/datduong/Classify-WS-22q-Img and https://github.com/datduong/stylegan2-ada-Ws-22q, respectively.

https://github.com/datduong/stylegan2-ada-Ws-22q

## Acknowledgments

This research was supported by the Intramural Research Program of the National Human Genome Research Institute, National Institutes of Health. This work utilized the computational resources of the NIH HPC Biowulf cluster (http://hpc.nih.gov).

## Figure Legends

**Figure 1**. Examples of GAN fake images for 22q and WS. Type 1 fake images (a) 22q (top row) and WS (bottom row) were generated with GAN and are all theoretically unique. Type 2 (top row) and type 3 (bottom row) fake images 22q (b) and WS (c) were generated using the same random vector in each age group. For type 2, general features, such as skin tone and hair color, are roughly preserved. For type 3, the images look consistent at depicting the same “person” progresses through different age groups. Type 4 fake images (d) were created images with blended facial characteristics of two disease labels. The main disease condition (22q or WS) represents 55% of the facial phenotype and the added disease condition represents 45% of the facial phenotype. For example, WS:Unaffected is a blend of 55% WS facial features and 45% unaffected facial features. Only the blended images were used for training, the left most images are shown here as references.

**Figure 2**. Confusion matrix of accuracy of classifier trained on real images. Rows represent the correct label, while columns represent the label chosen by the classifier. The diagonal numbers represent the percent accuracy for each category (the percentage of images when the correct label was identified), while the off-diagonal represent misclassification percentage ascribed to an incorrect category. Accuracy is based on 50 test images of WS, 50 of 22q and 81 of other conditions.

**Figure 3**. Occlusion analysis of 22q and WS facial images across the lifespan. Composite saliency maps (occlusion analysis) were generated by averaging all test images in each age groups: infant, child, adolescent, young adult, and older adult (reading left to right) for both 22q (a) and WS (b). Green and red indicate positive and negative contribution to the correct labels, respectively.

**Figure 4**. Rank of Euclidean distance matrix of key facial features during aging. Rank (in fraction) of observed Euclidean distance (out of 100 permutations) between embeddings of the averages of occlusion analysis for two age groups. A small number indicates key features identified by the neural network for two age groups are more statistically similar, whereas a larger number indicates key features are more statistically different.

