## Supplemental Table 1 for "Neural networks for classification and image generation of aging in genetic syndromes"

**Supplemental Table 1.** Description of genetic conditions studied.

| **Syndrome** | **Molecular Etiology** | **Inheritance pattern** | **Prevalence^a^** | **Common Features** | **Facial Features** | **Differential Diagnosis^b^** |
| --- | --- | --- | --- | --- | --- | --- |
| 22q11.2 Deletion Syndrome (other terms: DiGeorge syndrome, Velocardiofacial syndrome)^1^ | Heterozygous deletion of 22q11.2 (1.5 to 2.54 Mb)  (Genes of interest in deleted region: *TBX1*; *DGCR8*; *CRKL*; *SNAP29*; and *PRODH*) | de novo (~90%), Autosomal Dominant | 1/4500 to 1/10,000 at birth | Congenital heart defects (primarily conotruncal malformations); palatal anomalies; distinctive (but often subtle) facial features; hypocalcemia; immune deficiency; learning difficulties | Hypertelorism epicanthal folds; prominent nasal root; short philtrum; micronathia; low-set ears^2^ | Alagille syndrome; CHARGE syndrome;  Deletion 10p14-p14; Fetal alcohol spectrum; Goldenhar syndrome; Jacobsen syndrome; maternal diabetes, maternal retinoic acid exposure; Smith-Lemli-Opitz syndrome; VACTERL association |
| Williams Syndrome (other terms: Williams-Beuren syndrome)^3^ | Heterozygous deletion of 7q11.23 (1.55 to 1.83 Mb)  (Genes of interest in deleted region: *ELN*; *LIMK1*; *GTF2I*; *STX1A*; *BAZ1B*; *CLIP2*; *GTF2IRD1*; *NCF1*) | de novo (93%), Autosomal Dominant | 1/7500 | Elastin arteriopathy^4^ (supravalvular aortic stenosis, most common); distinctive facial features; intellectual disability; endocrine abnormalities; connective tissue abnormalities | Broad forehead; bitemporal narrowing; periorbital fullness; stellate irises; short, upturned nose; long philtrum; full lips^5^ | 22q11.2 Deletion syndrome; Fetal alcohol spectrum; Kabuki syndrome; Noonan syndrome; Smith-Magenis syndrome |

^a^ Orphanet. Prevalence and incidence of rare diseases: Bibliographic data, January 2019, Number 01. 2019. https://www.orpha.net/orphacom/cahiers/docs/GB/Prevalence_of_rare_diseases_by_diseases.pdf. Accessed 1 November 2021.  Prevalence is given for the population unless otherwise stated.

^b^The differential diagnoses for these conditions are based on the overall phenotype, not just the facial features.
