## Supplemental Table 2 for "Neural networks for classification and image generation of aging in genetic syndromes"

| ***Ancestry categories*** | ***Williams syndrome*** | ***22q11.2 deletion syndrome*** |
| --- | --- | --- |
| African and African American | 30 | 71 |
| European | 88 | 107 |
| Latino | 67 | 52 |
| Middle Eastern and Nord African | 23 | 10 |
| Asian | 73 | 58 |
| *Eastern Asian* | *18* | *40* |
| *Southeastern Asian* | *10* | *7* |
| *South Asian* | *40* | *10* |
| *Asian, unclassified* | *5* | *1* |
| Uncategorized* | 253 | 296 |
| *Total* | 534 | 594 |

* There was insufficient information to allow precise inclusion of images of these individuals in one of the 5 ancestry categories.

**Supplemental Table 2**: Ancestry distribution of all individuals with Williams syndrome and 22q11.2 deletion syndrome used in this study. Ancestry was recorded as reported on the image source (published articles or websites) and ancestry categories were defined by investigators based on available literature^1; 2^.

1. Huddart, R., Fohner, A.E., Whirl‐Carrillo, M., Wojcik, G.L., Gignoux, C.R., Popejoy, A.B., Bustamante, C.D., Altman, R.B., and Klein, T.E. (2019). Standardized biogeographic grouping system for annotating populations in pharmacogenetic research. Clinical Pharmacology & Therapeutics 105, 1256-1262.

2. Flanagin, A., Frey, T., Christiansen, S.L., and Bauchner, H. (2021). The reporting of race and ethnicity in medical and science journals: comments invited. JAMA 325, 1049-1052.
