## Supplemental Table 4 for "Neural networks for classification and image generation of aging in genetic syndromes"

|  | **Number of**  **Williams Syndrome Survey Participants** | **Accuracy of**  **Williams Syndrome Survey Participants** |  | **Number of**  **22q11.2 DS Survey Participants** | **Accuracy of**  **22q11.2 DS Survey Participants** |
| --- | --- | --- | --- | --- | --- |
| Total | 30 | 77.53% |  | 30 | 59.27% |
| **Experience** |  |  |  |  |  |
| < 5 years | 8 | 75.25% |  | 9 | 59.33% |
| 5 to 10 years | 7 | 76% |  | 6 | 60% |
| > 10 years | 15 | 79.47% |  | 15 | 58.93% |
| **Practice Setting** |  |  |  |  |  |
| Academic/Research | 25 | 78% |  | 25 | 58.96% |
| Other | 5 | 75.2% |  | 5 | 60.80% |
| **Geographic Practice Location** |  |  |  |  |  |
| Africa | 0 |  |  | 0 |  |
| Asia | 0 |  |  | 1 |  |
| Australia/Oceania | 0 |  |  | 0 |  |
| Europe | 0 |  |  | 0 |  |
| North America | 30 |  |  | 29 |  |
| South America | 0 |  |  | 0 |  |

Supplemental Table 4. Demographic Information and average accuracies for clinical geneticists who completed surveys.
