## Supplemental Table 5 for "Neural networks for classification and image generation of aging in genetic syndromes"

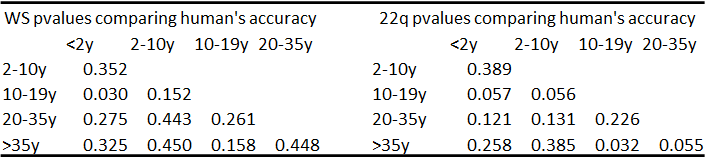

Supplemental Table 5. Comparing human’s performances among different age brackets. We averaged the accuracy for 10 test images in each age group (which are showed in Table 1), and then compare these averages between every pair of age groups. P-values were computed via permutation test for every age group pair (e.g., 2 sets of 10 images).
