## Supplemental Table 6 for "Neural networks for classification and image generation of aging in genetic syndromes"

|  | WS | Other Conditions for WS survey | 22q | Other Conditions for 22q survey | Other Conditions (both surveys combined) |
| --- | --- | --- | --- | --- | --- |
| Test Images Partial or Full Smile (n) | 30/50 (60%) | 22/50 (44%) | 12/50 (24%) | 20/50 (40%) | 34/81  (42%) |
| Partial Smile (n) | 6 | 9 | 8 | 8 | 14 |
| Full Smile (n) | 24 | 13 | 4 | 12 | 20 |
| Overall Accuracy | 72.80% | 82.27% | 57.20% | 61.33% | 71.38% |
| Smile (Partial and Full) | 82.44% | 80.30% | 53.89% | 58.67% | 70.39% |
| Full Smile | 85.56% | 79.48% | 46.67% | 57.33% | 69.00% |
| Partial Smile | 70% | 81.48% | 57.50% | 59.17% | 72.38% |
| No Smile | 58.33% | 83.81% | 58.24% | 63.11% | 72.10% |

Supplemental Table 6. Clinical Geneticist accuracy in smiling vs. non-smiling test images in across both surveys. If an “other conditions” image was shown in both surveys, the average accuracy was calculated and used to determine the overall average.
