## Supplemental Table 7 for "Neural networks for classification and image generation of aging in genetic syndromes"

|  | **Number of**  **Williams Syndrome Survey Participants** | **Accuracy of**  **Williams Syndrome Survey Participants** |  | **Number of**  **22q11.2 DS Survey Participants** | **Accuracy of**  **22q11.2 DS Survey Participants** |
| --- | --- | --- | --- | --- | --- |
| Total | 30 | 77.53% |  | 30 | 59.27% |
| **Consideration of Facial Features in Diagnosis** |  |  |  |  |  |
| Major factor | 15 | 79.07% |  | 3 | 65.33% |
| Intermediate factor | 12 | 76.33% |  | 19 | 61.15% |
| Minor factor | 3 | 74.67% |  | 8 | 52.5% |
| Not used in diagnosis | 0 | - |  | 0 | - |
| **Age Easiest to Accurately Diagnose Based on Facial Features** |  |  |  |  |  |
| Infants (< 2 y/o) | 0 | - |  | 0 | - |
| Early Childhood (2 to 9 y/o) | 27 | 76.59% |  | 24 | 59.33% |
| Adolescence (10 to 19 y/o) | 1 | 74% |  | 4 | 61% |
| Younger Adults (20 to 35 y/o) | 1 | 88% |  | 2 | 55% |
| Older Adults (> 35 y/o) | 1 | 96% |  | 0 | - |
| **Age Most Difficult to Accurately Diagnose Based on Facial Features** |  |  |  |  |  |
| Infants (< 2 y/o) | 14 | 78.42% |  | 18 | 59.67% |
| Early Childhood (2 to 9 y/o) | 0 | - |  | 0 | - |
| Adolescence (10 to 19 y/o) | 0 | - |  | 0 | - |
| Younger Adults (20 to 35 y/o) | 1 | 76% |  | 0 | - |
| Older Adults (> 35 y/o) | 15 | 76.8 |  | 12 | 58.67% |
| **Influence of Ancestral Origin on Presentation of Characteristic Facial Features** |  |  |  |  |  |
| Greatly influences presentation | 4 | 76% |  | 9 | 58.22% |
| Some influence on presentation | 22 | 74% |  | 19 | 60.10% |
| Little to no influence on presentation | 4 | 82.5% |  | 2 | 56% |

**Supplemental Table 7.** Attitudes and opinions of survey participants compared to their overall accuracy.
