## Supplemental Table 8 for "Neural networks for classification and image generation of aging in genetic syndromes"

| Condition | Infant/  < 2 y/o | Child  2 to 9 y/o | Adolescent  10 to 19 y/o | Young Adult 20 to 34 y/o | Older Adult  ≥35 y/o | Overall Average |
| --- | --- | --- | --- | --- | --- | --- |
| 22q11.2 Deletion Syndrome | 0.88 y/o  (2.12 y/o) | 5.54 y/o  (7.75 y/o) | 13.44 y/o  (17.49 y/o) | 25.40 y/o  (27.83 y/o) | 43.51 y/o  (43.21 y/o) | 11.73 y/o (13.98 y/o) |
| Williams  Syndrome | 0.91 y/o  (1.54 y/o) | 5.01 y/o  (6.21 y/o) | 13.97 y/o  (16.84 y/o) | 25.40 y/o  (27.80 y/o) | 48.92 y/o  (50.00 y/o) | 11.94 y/o (13.52 y/o) |
| Other Conditions | 0.73 y/o  (2.42 y/o) | 5.33 y/o  (7.13 y/o) | 13.83 y/o  (16.72 y/o) | 24.88 y/o  (27.54 y/o) | 46.13 y/o  (44.29 y/o) | 14.05 y/o (15.86 y/o) |

**Supplemental Table 8.** Average age for each condition.

An average age for individuals shown in training and test set images. Ages were initially obtained through published content. If age was not given, the research team, at least 2 researchers per individual image, determined an empiric age for the individual. Additionally, age of individuals was derived using the FairFace age classifier, which was trained on presumably unaffected, healthy individuals. These derived values are shown in parentheses.
