## Supplemental Figure 1 for "Neural networks for classification and image generation of aging in genetic syndromes"

### Slide 1
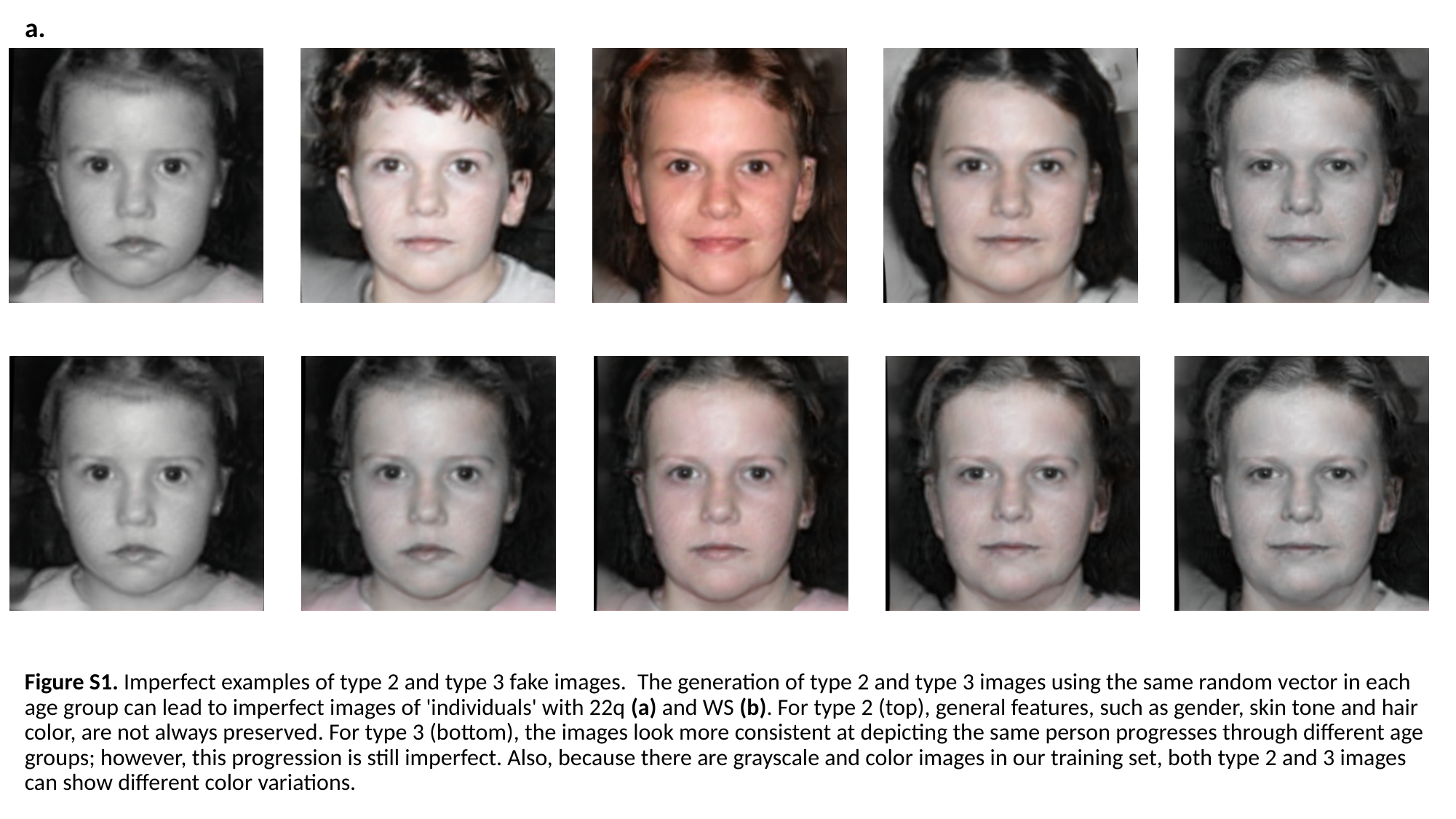

a.
Figure S1. Imperfect examples of type 2 and type 3 fake images.  The generation of type 2 and type 3 images using the same random vector in each age group can lead to imperfect images of 'individuals' with 22q (a) and WS (b). For type 2 (top), general features, such as gender, skin tone and hair color, are not always preserved. For type 3 (bottom), the images look more consistent at depicting the same person progresses through different age groups; however, this progression is still imperfect. Also, because there are grayscale and color images in our training set, both type 2 and 3 images can show different color variations.

### Slide 2
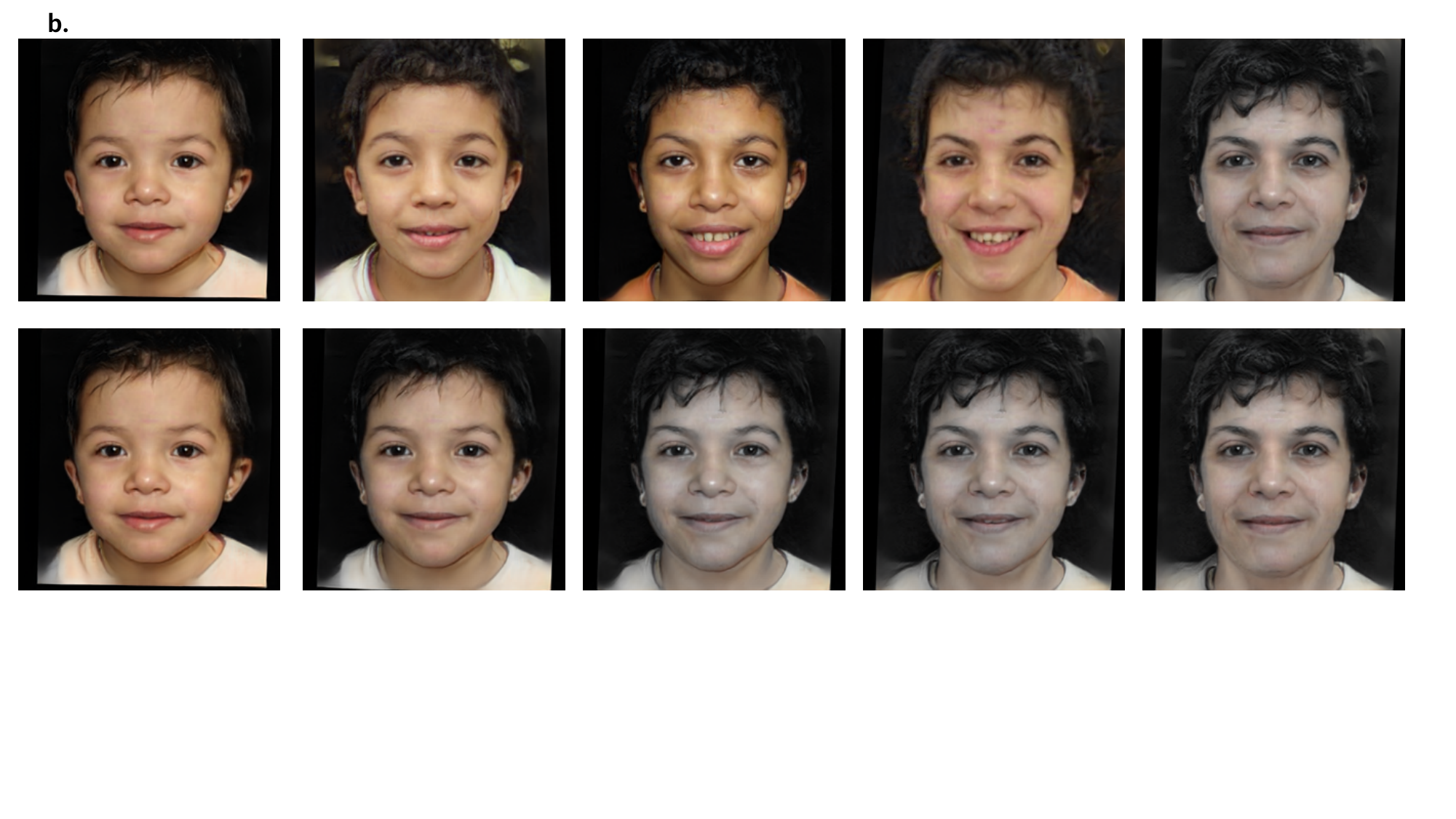

b.
