## Supplemental Table 3 for "Neural networks for classification and image generation of aging in genetic syndromes"

| Condition | Infant/  < 2 y/o | Child  2 to 9 y/o | Adolescent  10 to 19 y/o | Young Adult 20 to 34 y/o | Older Adult  ≥35 y/o | Total |
| --- | --- | --- | --- | --- | --- | --- |
| 22q11.2 deletion syndrome | 95 | 236 | 98 | 90 | 25 | 544 Training Images |
|  | 10 | 10 | 10 | 10 | 10 | 50 Test Images |
| Williams  syndrome | 85 | 211 | 101 | 58 | 29 | 484 Training Images |
|  | 10 | 10 | 10 | 10 | 10 | 50 Test Images |
| Other  conditions | 96 | 260 | 149 | 115 | 65 | 685 Training Images |
|  | 14* | 19* | 18* | 17* | 13* | 81* Test Images |
| All | 310 | 746 | 386 | 300 | 152 | 1894 |

**Supplemental Table 3.** Number of images for each age grouping. In addition to training set images (top number), ten unique images were used to the training images to test the model. *For other conditions two sets of 10 images were used in the test set, some of the same images were used for both the 22q11.2 deletion syndrome and Williams syndrome test. Confusion matrices were made based on the 50 Williams syndrome images, 50 22q11.2 deletion syndrome images, and 81 other conditions images.
